# Shared Strides: Operational feasibility of community-based biomechanics data collection in knee osteoarthritis

**DOI:** 10.64898/2026.04.20.26351135

**Authors:** Ryan C. McCloskey, Jenna M. Qualter, Alex Gruber, Sean Leapley, Peihua Qiu, Zibo Tian, Heather K. Vincent, Kerry E. Costello

**Author notes:** **Corresponding Author Address**: Mechanical & Aerospace Engineering, University of Florida Attn: Kerry Costello, 1064 Center Drive, Building NEB Room 181 – Costello Lab Gainesville, FL 32611, USA. **E-mail addresses:**.

## Abstract

Biomechanics studies using traditional optical motion capture have been limited by small, homogeneous sample sizes and a focus on single movements, restricting the ability to capture clinically relevant adaptations across daily tasks. These limitations are particularly consequential in heterogeneous musculoskeletal conditions such as knee osteoarthritis (OA), where variability in demographic and clinical characteristics necessitates large, representative samples to identify patient-specific biomechanical intervention targets. Markerless motion capture enables faster, high-throughput data collection and offers the potential for community-based assessments; however, its feasibility of use in clinical populations across diverse tasks remains unclear. This study evaluated the feasibility of community-based, high-throughput markerless biomechanics data collection in individuals with knee OA. Participants (n = 85) completed a series of activities of daily living using a portable markerless motion capture system deployed across two community-based and two on-campus sites. Feasibility was assessed using timing metrics related to research operations (transit, setup, calibration, breakdown), participant workflow (consent, questionnaires, motion capture), and task-specific durations. No significant differences in timing metrics were observed across sites despite logistical and operational challenges. These findings support the feasibility of using high-throughput, community-based markerless motion capture and suggest a viable pathway for addressing long-standing limitations in sample size and representativeness through scalable data collection workflows in biomechanics studies.

## 1. Introduction

In musculoskeletal conditions characterized by substantial inter-individual variability, such as knee osteoarthritis (OA) (Deveza and Loeser, 2018; White et al., 2016), biomechanical contributors to pain and dysfunction are likely to be patient-specific. Identifying these biomechanical contributors and potential intervention targets, therefore, requires studies with sufficiently large, clinically representative samples and assessment across a range of functional movements.

Prior work in OA has identified associations between gait metrics, such as the knee adduction moment (KAM), and long-term cartilage degeneration (Bennell et al., 2011; Chang et al., 2015). Other common activities of daily living (ADLs), including stair climbing, sit-to-stand transitions, and squatting, have also provided important insights into knee OA progression, as these tasks place unique demands on the knee and may reveal pathological loading patterns not evident during level walking (Amin et al., 2008; Gonçalves et al., 2017; Verlaan et al., 2018). However, these insights have largely been derived from studies focused on isolated movements (Amin et al., 2008; Gonçalves et al., 2017; Verlaan et al., 2018) and relatively small, homogeneous samples (Ory et al., 2002; Vagenas et al., 2018), which may contribute to variability in response to biomechanical interventions across individuals (Favre et al., 2016; Rynne et al., 2022) and limit the ability to identify patient-specific, disease-relevant loading patterns across diverse functional contexts.

A major contributor to these longstanding limitations is the reliance on marker-based optical motion capture. While widely used for assessing movement biomechanics (Das et al., 2023), these systems are typically constrained to laboratory settings, require highly trained personnel to operate, and result in time-intensive data collections (Chen et al., 2016; Gorton et al., 2009; Robles-García et al., 2015; Simon, 2004). Markerless motion capture provides a promising alternative, using deep learning-based pose estimation to infer body segment positions from high-speed video. This technology eliminates the need for physical markers and specific clothing requirements, reducing per-participant data collection time and enabling a high-throughput data collection model. This model can facilitate off-site data collection in community settings (e.g., clinics, gyms, assisted living communities) (McGuirk et al., 2022), which may lower barriers to participation and increase the heterogeneity of recruited populations (Rigatti et al., 2022; Samus et al., 2015; Wieland et al., 2021). Together, these advances provide a pathway toward more representative, large-scale biomechanical datasets needed to understand variability in disease progression and support patient-specific intervention strategies.

McGuirk et al. (2022) demonstrated the preliminary feasibility of a high-throughput markerless motion capture model across six community sites. However, this work did not include a clinical population, and assessments were limited to walking tasks. A significant evidence gap remains regarding the feasibility of extending high-throughput markerless motion capture to clinical populations and a broader range of functional tasks. Therefore, the primary objective of this study was to compare data collection timing metrics between on-campus and community-based settings during high-throughput assessment of ADLs in individuals with knee OA.

## 2. Methods

### 2.1. Shared Strides Study Overview

The Shared Strides study was designed to assess the feasibility of community-based biomechanics data collection in individuals with knee OA. Data collections were performed as high-throughput, single-day data collection “events” at four sites: 1) an event room in the central building of a gated residential senior living community (Retirement Community), 2) a classroom within a public day-use senior community recreation center (Senior Center), 3) a laboratory located behind a physical therapy clinic at a quaternary care medical center (Clinical Laboratory), and 4) an event room on the lower-level of a university student union (Student Union). Twelve data collections (eleven full day, one half day) were conducted over an 11-month period (Table 1).

**Table 1.** Data collection site information.

| Site | Retirement Community | Senior Center | Clinical Laboratory | Student Union |
| --- | --- | --- | --- | --- |
| Distance from motion analysis lab | 4.1 miles | 6.3 miles | 2.9 miles | 600 ft |
| n, completed data collection dates | 3* | 4 | 3 | 2 |

\*One collection was a half day

A portable markerless motion capture system was transported to and from site locations using a university-owned pick-up truck equipped with a lift gate (Fig. 1B). Equipment was organized into a hard case containing force plates, a custom-built mobile computer cart, and two additional carts containing the remaining equipment and supplies (Fig. 1A). The Student Union site was adjacent to the motion analysis lab where the equipment was stored when not in use, and did not require vehicular transport, therefore all equipment was carried on foot.

**Figure 1.**
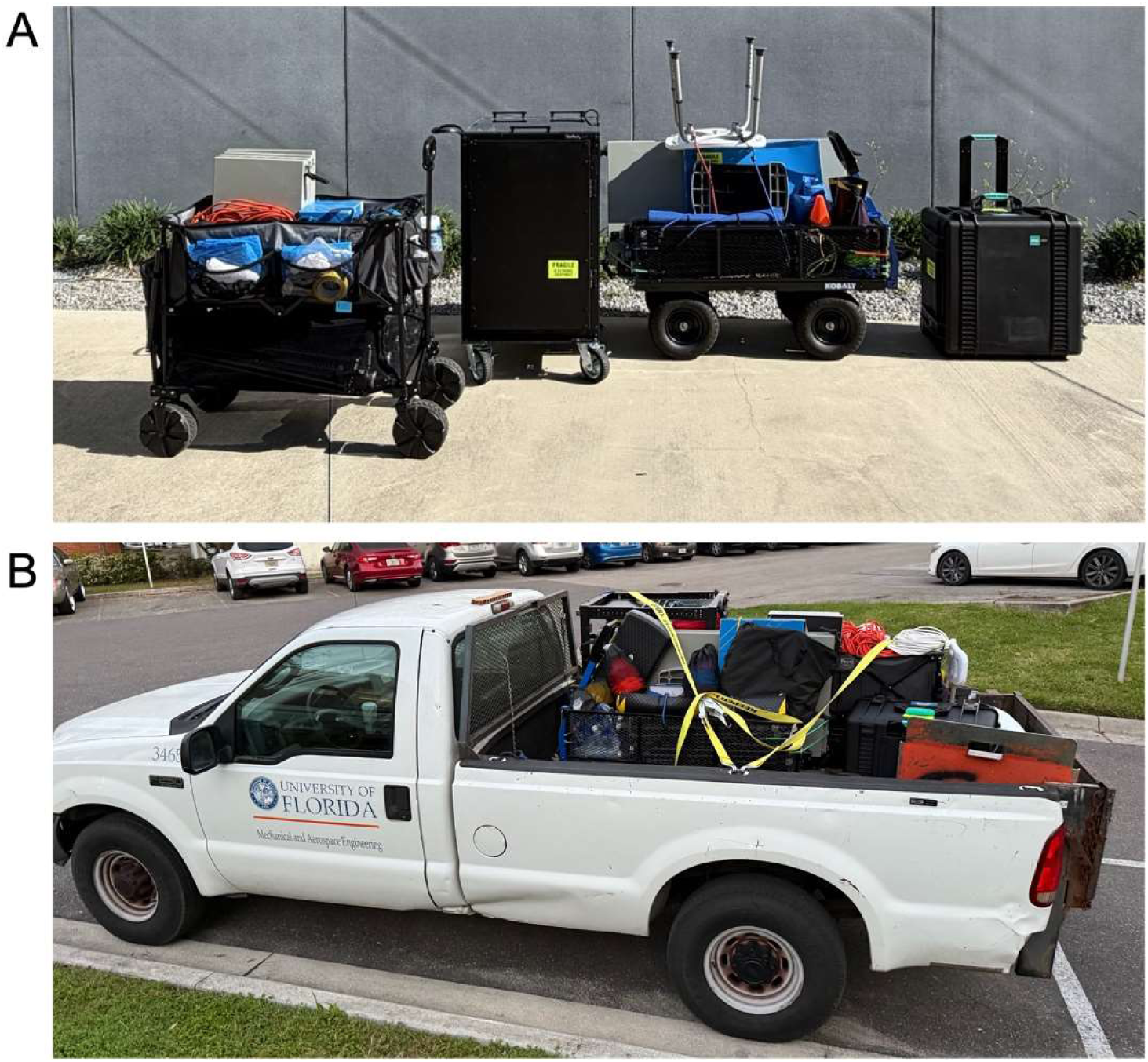
A) Packed markerless motion capture system B) Portable system in transport truck.

Data were collected from multiple participants per day, with each completing informed consent, questionnaires, and a motion capture assessment consisting of a set of ADLs. Due to the nature of the high-throughput model, the order of questionnaires and motion capture varied depending on scheduling and equipment availability. Participants returned for a follow-up session six months after their initial visit. Some dates included a mixture of initial and follow-up visits. The current analysis includes data from initial visits only.

All procedures were approved by the University of Florida Institutional Review Board (IRB202400674), and all participants provided written informed consent before participating in any study procedures.

### 2.2. Study Sample

To determine eligibility, participants were either pre-screened by phone or screened on site as “walk-ins.” Pre-screened individuals scheduled a data collection appointment, whereas walk-in participants either completed data collection on the same day or were scheduled for a later date. Eligible participants were adults aged 40 or older with a self-reported diagnosis of knee OA in one or both knees, or those meeting the American College of Rheumatology (ACR) clinical criteria (Altman et al., 1986) in one or both knees. Additionally, participants were required to be able to walk 20 feet without assistance. Exclusion criteria included inability to provide informed consent, inability to read or follow directions in English, self-reported pregnancy, lower limb injury or surgery within the past 3 months, joint replacement surgery in the affected knee(s), or other self-reported conditions (e.g., stroke, Parkinson’s disease, degenerative nerve conditions, acute spine pain, or current cancer treatment) that affect movement more than the diagnosis of OA.

### 2.3. Questionnaires and Anthropometrics

Participants electronically completed a set of questionnaires in REDCap (Research Electronic Data Capture) using a study-owned iPad: custom demographics and acceptability questionnaires, the short-form International Physical Activity Questionnaire (IPAQ), and a custom clinical history questionnaire primarily focused on knee joint health. Participants also completed standardized symptom questionnaires (the Knee Injury and Osteoarthritis Outcome Scores [KOOS] and the Intermittent and Constant Osteoarthritis Pain [ICOAP] scale) for each knee with OA (left, right, or both). Questionnaire start and end times were recorded.

Height and weight were measured by study personnel for each participant and used to calculate body mass index (BMI).

### 2.4. Markerless Motion Capture

The markerless motion capture setup utilized eight FLIR (Blackfly S, Teledyne FLIR Systems, USA) cameras (100 Hz), and two Vicon Vero (Vicon, Oxford, UK) cameras (100 Hz), mounted on tripods in an oval pattern to define the capture volume (Fig. 2). The capture area was centered around a 3.6 m Accugait walkway, with two embedded AMTI (ACG-O, AMTI, MA, USA) force platforms (1000 Hz).

**Figure 2.**
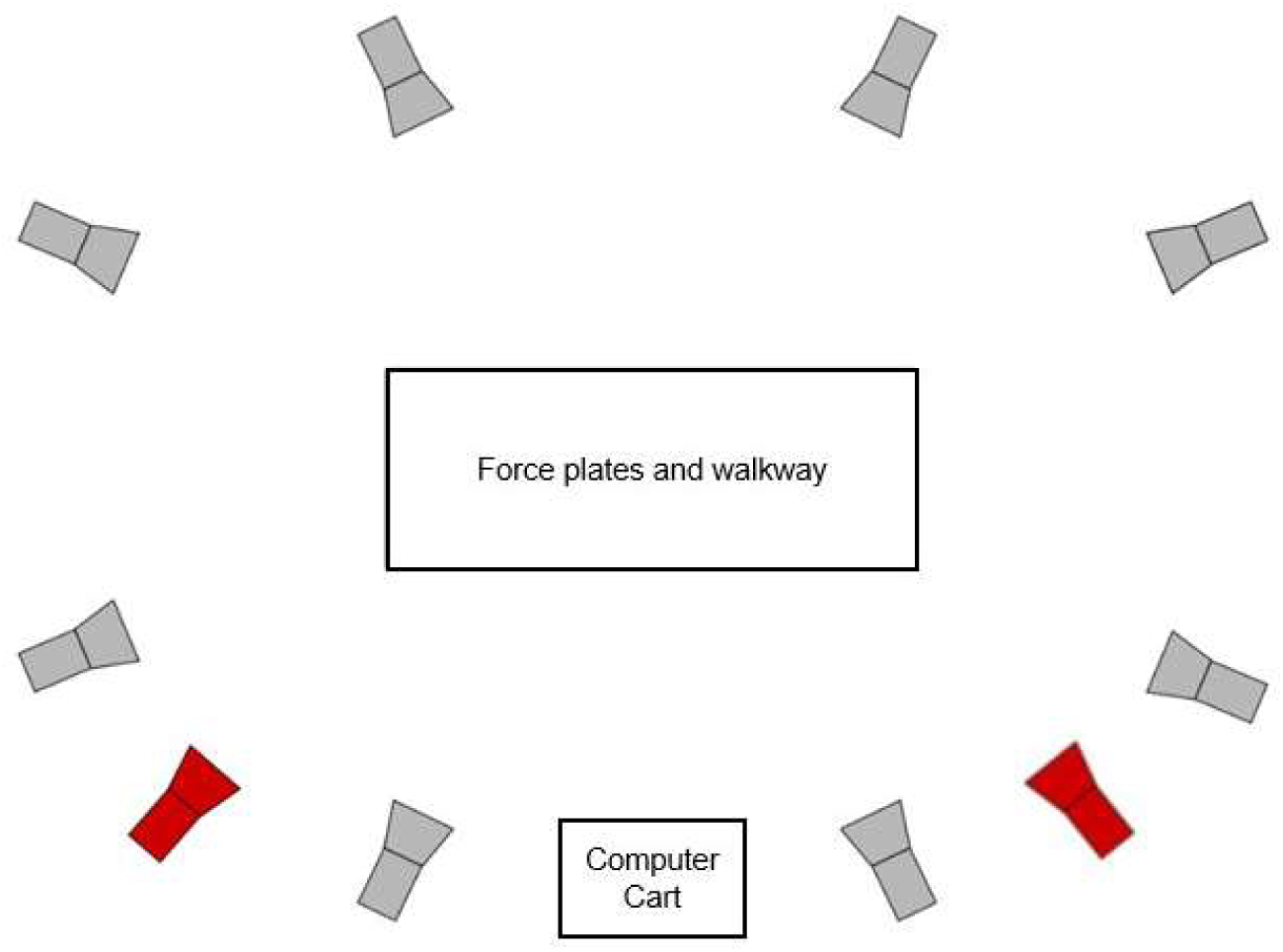
Markerless motion capture set-up schematic (FLIR cameras in grey; Vicon Vero cameras in red)

Standard setup and collection protocols (i.e., consistent tripod height and layout) were followed at all sites to ensure consistency in data collection procedures. All data were recorded through Vicon Nexus software and stored on PC hard drives for later processing with Theia3D software (Theia Markerless Inc., ON, Canada).

The motion capture portion of the study involved a set of ADLs, performed within the capture volume under the direction of a study team member. These included self-selected and fast-paced overground walking, double- and single-leg balance, squatting, sit-to-stand, and step-up/down (Appendix A). Any activity could be omitted due to participant preference (e.g., due to discomfort or inexperience with the task) or at the discretion of the study staff if safety concerns arose (e.g., due to fall risk). In some cases, participants attempted a practice trial or failed trial(s) (e.g., unable to keep one leg off the floor during single-leg balance) before deciding to skip the activity. The goal was to collect at least three successful trials per activity, except for the double-leg standing balance, which was performed as a single 10-second trial. Total trial counts were recorded for each activity.

### 2.5. Data Analysis

Feasibility was assessed using timing metrics related to research operations (study team transit, setup, calibration, breakdown) and participant workflow (informed consent, questionnaires, motion capture) at each site. Time durations for each segment and activity were calculated from recorded timestamps and rounded to the nearest minute, with durations shorter than one minute rounded up. Timestamps were recorded by study personnel using synchronized digital devices (e.g., smartphones). Total participant time was defined as the duration from initial walk-in to completion of all questionnaires and motion capture tasks, including any idle time waiting for equipment to become available. The number of research team members present during setup and breakdown was also documented at each site.

Differences in participant timing metrics across sites were assessed using Welch’s one-way analysis of variance (ANOVA) with significance set at α = 0.05. Research operations differences were reported descriptively only due to the small number of dates per site.

## 3. Results

Across the four sites, 85 participants completed initial visits (Table 2). Site-level data are not specific to visit type; thus, site data from all dates/sites with initial visits are included.

**Table 2.** Participant population characteristics by site.

| Site | Retirement Community | Senior Center | Clinical Laboratory | Student Union |
| --- | --- | --- | --- | --- |
| **n, participants** | 26 | 39 | 13 | 7 |
| **Sex, n (%) Female** | 19 (73.1%)* | 30 (76.9%) | 10 (76.9%) | 4 (57.1%) |
| **Age** | 72.8 ± 12.0 | 73.1 ± 7.3 | 73.8 ± 5.5 | 71.7 ± 8.7 |
| **BMI** | 28.7 ± 6.9 | 27.8 ± 5.9 | 29.0 ± 6.3 | 26.1 ± 3.1 |
| **KOOS Pain\*\*** | 69.3 ± 18.8 | 64.3 ± 16.1 | 69.6 ± 21.4 | 71.7 ± 17.4 |

\*One participant selected "Prefer not to say" for sex.
\*\*KOOS scores are reported on a 0-100 scale for the knee with highest pain; lower scores indicate greater pain.

### 3.1. Site-Level Timing

The markerless motion capture system was successfully transferred from the motion analysis lab to each study site (Table 3). While the limited number of dates per site did not allow formal statistical analysis of timing differences in research operations across sites, visual observation suggests setup and breakdown times were similar across sites, with some reductions in setup/breakdown time with more staff assistance. Community-based collections exhibited longer setup times on average than on-campus locations (35.3 minutes vs 26.6 minutes; Table 3).

**Table 3.** Implementation characteristics per data collection.

| Mo. | Site | n, participants | Transit time (minutes) | Setup time (minutes) | n, staff (setup) | Calibration time (minutes) | Break down time (minutes) | n, staff (break down) |
| --- | --- | --- | --- | --- | --- | --- | --- | --- |
| 1 | Senior Center | 8 | 20 | 48 | 4 | 39 | 29 | 7 |
| 3 | Clinical Laboratory | 6 | X | 27 | 5 | 29 | 30 | 4 |
| 3 | Senior Center | 13 | 37 | 30 | 4 | 29 | X | 7 |
| 6 | **Retirement Community** | 14 | 18 | 27 | 7 | 65 | 49 | 4 |
| 6 | **Student Union** | 6 | 2 | 26 | 6 | 11 | 20 | 8 |
| 7 | **Clinical Laboratory** | 4 | 17 | 25 | 8 | 25 | 50 | 5 |
| 8 | **Senior Center** | 4 | 25 | 36 | 5 | 82 | 38 | 5 |
| 9 | **Clinical Laboratory** | 3 | 11 | 29 | 7 | 170 | 25 | 5 |
| 11 | **Senior Center** | 14 | 18 | 35 | 6 | 65 | 23 | 6 |
| 11 | **Student Union** | 1 | 6 | 26 | 6 | 15 | 27 | 5 |
| 12 | **Retirement Community** | 6 | 22 | 34 | 4 | 31 | 25 | 6 |
| 12 | **Retirement Community** | 6 | 17 | 37 | 5 | 21 | 29 | 4 |

X indicates missing data Abbreviations: Mo. = month

### 3.2. Participant-Level Timing

Mean data collection durations for informed consent, questionnaires, and motion capture assessments were not statistically different across the four sites, with site-level mean total visit time varying by only 2.3 minutes (Table 4). Slight data losses occurred across all sites due to missing start or end timestamps (<1%) or not completing all segments of the data collection protocol (1 participant had to leave early and did not complete motion capture; data are included for consent, questionnaire, and total visit time).

**Table 4.** Timing metrics per site (mean ± SD). All times are reported in minutes.

| **Data Collection Segment** | **Retirement Community** | **Senior Center** | **Clinical Laboratory** | **Student Union** | **p-value** |
| --- | --- | --- | --- | --- | --- |
| **Informed Consent** | 11.0 ± 5.5<br>(n=24) | 10.4 ± 2.4<br>(n=37) | 11.5 ± 5.5<br>(n=11) | 9.7 ± 1.3<br>(n=7) | 0.495 |
| **Questionnaires** | 22.4 ± 7.9<br>(n=25) | 26.8 ± 10.5<br>(n=38) | 26.6 ± 6.3<br>(n=13) | 27.2 ± 3.1<br>(n=6) | 0.130 |
| **Motion Capture** | 22.0 ± 4.3<br>(n=25) | 21.6 ± 4.5<br>(n=38) | 25.5 ± 4.9<br>(n=13) | 24.1 ± 3.9<br>(n=7) | 0.085 |
| **Total Visit\*** | 66.4 ± 20.4<br>(n=26) | 65.1 ± 15.7<br>(n=37) | 67.3 ± 10.3<br>(n=13) | 65.6 ± 6.9<br>(n=7) | 0.946 |

\*Total visit time includes time spent waiting between data segments

The time required to complete individual ADLs ranged from 1 to 9 minutes, with variability across activities (APPENDIX B). Sixty-nine participants (81.2%) completed all ADLs. Single-leg balance and step-up/down trials were the most frequently skipped. Walking trials exhibited the longest durations and greatest variability among ADLs (4.2 ± 1.7 minutes for self-selected pace; 3.7 ± 1.2 for fast pace); successful trials required clean foot strikes on the walkway-embedded force plates.

## 4. Discussion

The findings from this study demonstrate that high-throughput, community-based markerless motion capture assessments of ADLs are feasible in individuals with knee OA. Over 12 data collection events, biomechanics data across a comprehensive set of ADLs were collected from 85 participants, with up to 18 participants assessed in a single day. Sample sizes of this magnitude are often difficult to obtain in traditional biomechanics research; thus, these results support the potential for this approach to improve the scalability and efficiency of biomechanical data collection in clinical populations.

The average data collection duration of approximately 66 minutes per participant in the current study was longer than the 20 minutes per participant previously reported by McGuirk et al. (McGuirk et al., 2022). This was expected given the differences in study protocols. While McGuirk et al. measured regular and fast-paced walking across several passes (6-8) on a pressure mat and administered two questionnaires (McGuirk et al., 2022), the current study included a broader set of ADLs with six questionnaires. Importantly, activities were completed using a portable force plate-embedded walkway to capture kinetic data, requiring repeated trials to obtain sufficient clean force plate strikes. Thus, the approximate 45-minute difference in per-participant data collection reflects an intentional trade-off between session duration and the acquisition of more comprehensive biomechanical and patient-reported data. Importantly, the duration of the current protocol still falls below that of traditional marker-based motion capture assessments, which typically last approximately 1.5 to 2 hours in laboratory environments (Simon, 2004). Additionally, the high-throughput design allows for concurrent participation, such that multiple participants can complete different components of the study simultaneously, increasing the total number of participants assessed per day. Population differences may also have contributed to the longer data collection time in the current study, as both OA (Favre et al., 2014; Mills et al., 2013) and older age (Favre et al., 2014; Kerrigan et al., 1998) are associated with slower completion of activities such as walking.

Although not statistically different, a trend toward differences in motion capture time was observed between sites (p = 0.085). This may partially reflect site-specific workflow demands. For example, the Clinical Laboratory, which typically had fewer participants per day, may have required less emphasis on efficiency compared to higher-throughput settings. Differences in the number of skipped activities per site may have also contributed to this trend, however, anecdotally, individuals with higher function or lower pain often completed the activities more quickly than those who ultimately skipped activities. This variability reflects heterogeneity in functional capacity among individuals with knee OA and underscores the value of comprehensive assessment across a wide range of functional movements. Importantly, the observed difference in motion capture time between sites was relatively small (approximately 4 minutes), suggesting that, despite this trend, the practical impact of site-related differences is likely minimal.

Participants in this study were allowed to self-select data collection sites and available dates, resulting in higher participant numbers at community-based sites (n = 65) than on-campus sites (n = 20). This pattern is consistent with our corresponding assessment of participant acceptability, which indicated that community sites attracted a higher number of walk-in participants (n = 34) and first-time research participants, reflecting greater engagement in these settings (Qualter et al., pre-print).

Higher enrollment and more walk-ins at community sites could reasonably be expected to challenge workflow efficiency and increase participant idle time. However, the average idle time across all sites, calculated as the difference between average total visit time and the sum of average component times, was only 7.1 minutes. This low idle time reflects a well-coordinated workflow in which study personnel could adapt the order of surveys and motion capture assessments in real time (e.g., pausing questionnaires to complete motion capture assessments) to prevent bottlenecks and minimize downtime. This flexibility is a critical factor in sustaining a high-throughput data collection model and supporting participant retention. Prior work in the healthcare field has shown that operational challenges such as participant arrival variability (e.g., late arrivals, no-shows, rescheduling) and extended wait times negatively affect perceived quality of care, confidence in healthcare providers, and willingness to return for follow-up visits (Alrasheedi et al., 2019; Camacho et al., 2006; Elemoah et al., 2025; Nyce et al., 2021). In the current study, the low idle time suggests that community-based markerless motion capture assessments can be operationally efficient without compromising participant experience. Moreover, the higher number of walk-in participants at the community sites further supports the potential for community-based settings to increase engagement in biomechanics research, consistent with prior work in other fields (Rigatti et al., 2022; Samus et al., 2015; Wieland et al., 2021).

Aligned with logistical requirements identified in previous studies (e.g., ensuring accessible power sources and sufficient lighting (McGuirk et al., 2022)), all sites met these criteria prior to scheduling data collection. Site setup and breakdown times, which varied slightly depending on the number of research team members present, averaged approximately 30 minutes. While this represents a rate-limiting step in the data collection workflow, a single setup was sufficient for all participant assessments conducted during each event. In contrast, variability in calibration times was primarily driven by limitations inherent to the markerless motion capture system, compounded by the limited availability of remote technical support due to time zone differences. These factors occasionally reduced opportunities for troubleshooting and increased participant wait times, reflecting practical challenges of multi-site implementation that should be considered in future deployment of high-throughput workflows.

While the feasibility of data collection was demonstrated across sites, several limitations should be acknowledged. First, the interpretation of ADL completion times was limited by participant-specific variations in protocol. For example, some participants skipped activities entirely; others initially attempted activities but did not complete them; and some participants required more rest between activities than others. As a result, these variations may have influenced the recorded duration of preceding or adjacent tasks. Finally, total visit time should be interpreted in the context of minor differences in data collection workflows. Although procedures were consistent across sites, the recorded end time occasionally encompassed post-collection activities (e.g., compensation or follow-up scheduling) rather than the final data collection task.

Consequently, total visit duration may modestly overestimate active data collection time for some participants. Despite these limitations, the detailed characterization of timing in this high-throughput data collection model provides practical insight into workflow variability and visit structure, which could inform the design and implementation of community-based biomechanics studies, particularly in clinical populations such as knee OA.

## 5. Conclusion

This study demonstrates that high-throughput markerless motion capture to assess a broad range of ADLs can be feasibly implemented across campus and community-based settings in individuals with knee OA, with consistent performance across sites despite occasional technical challenges. The ability to efficiently collect full kinematic and kinetic data across diverse settings in clinical populations provides a pathway toward larger, more representative biomechanical datasets, addressing longstanding challenges in sample size and representativeness in traditional laboratory-based studies. As this approach continues to evolve, future work should evaluate data quality and participant retention across sites, as well as opportunities to optimize study design (e.g., team size, ADL selection) for specific research questions or populations.

Ultimately, integration of markerless motion capture into community settings may support more scalable, person-centered biomechanics research and inform data-driven, personalized approaches to care.

## Data Availability

All data produced in the present study are available upon reasonable request to the authors

## CRediT authorship contribution statement

**Ryan C. McCloskey:** Writing – original draft, Visualization, Methodology, Data curation, Formal analysis. **Jenna M. Qualter:** Writing – reviewing & editing, Visualization, Methodology, Data curation, Formal analysis. **Alex Gruber:** Writing – reviewing & editing, Methodology, Data curation. **Sean Leapley:** Writing – reviewing & editing, Data curation, Formal analysis. **Peihua Qiu:** Writing – reviewing & editing, Formal analysis. **Zibo Tian:** Writing – reviewing & editing, Formal analysis. **Heather K. Vincent:** Writing – reviewing & editing, Methodology. **Kerry E. Costello:** Writing – review & editing, Supervision, Resources, Project administration, Methodology, Funding acquisition, Formal analysis, Conceptualization.

## Declaration of competing interests

The authors declare that they have no known competing financial interests or personal relationships that could have appeared to influence the work reported in this paper.

## Funding

This research was supported by a grant from the National Institute on Aging, Claude D. Pepper Older Americans Independence Center at the University of Florida (P30 AG028740). This project utilized REDCap, which is supported by the National Center for Advancing Translational Sciences of the National Institutes of Health (UL1TR001427).

KEC was supported by an Investigator Award from the Rheumatology Research Foundation. RCM was supported by a Herbert Wertheim College of Engineering Dean’s Research Award from the University of Florida.

## Declaration of generative AI and AI-assisted technologies in the manuscript preparation process

During the preparation of this work the author(s) used ChatGPT (GPT-5.3 and earlier models; OpenAI) during final manuscript editing in order to improve language clarity and flow. After using this tool/service, the author(s) reviewed and edited the content as needed and take(s) full responsibility for the content of the published article.

## Ethics Statement

This study received ethical approval from the University of Florida IRB (IRB202400674) on October 4, 2024.

## Acknowledgements

We thank the study participants for contributing their time and the staff at the community sites for allowing use of their spaces and assisting with room scheduling and on-site logistics.

We acknowledge the contributions of over 25 AI Biomechanics Laboratory students who assisted with study piloting, logistics, and data collection, in particular graduate students Alexandra Chertok, Peter Schaefer, and Kaitlin Southern. We also thank Kaitlyn Downer and Colin Larke from collaborating laboratories for assisting with data collection during summer sessions.

We acknowledge Clinical Research Core support from the Claude D. Pepper Older Americans Independence Center at the University of Florida, including early guidance on study design and regulatory preparation from Dr. Stephen Anton, study coordination (including recruitment outreach, participant screening, and reminder calls) from Maryam Sohooli, and early coordination support from Nevena Stanojevic.

We also acknowledge Austin Leto for his contributions to the design of the mobile PC system, including coordination of hardware and software requirements to support portable data collection and storage workflows.

## Appendix A: Descriptions of each activity of daily living task included in the movement assessment for this study

**Representative images of the activities are not included to preserve participant anonymity but are available from the corresponding author upon reasonable request.**

### Walking Tasks

Participants completed 2 walking conditions: self-selected pace and fast pace. For each condition, participants were instructed to walk at their own pace across the capture volume, starting and ending between two sets of small cones two meters beyond each side of the walkway. They were instructed to swing their arms naturally at their sides and to keep their gaze focused on a point directly ahead at eye level. For fast-paced walking, participants were instructed to walk as fast as they could safely go without running. A successful trial was defined as one in which at least one foot landed fully within a force plate’s perimeter. To achieve the desired number of strikes, starting cone positions were adjusted between trials.

### Balance Tasks

Participants performed both double-leg and single-leg balance trials. For double-leg balance, a 10-second trial was recorded with each foot fully on separate force plates. For single-leg balance, 5-second trials were recorded per leg on a single force plate. Before recording, participants were asked whether they could balance on one leg for 5 seconds. Those who indicated they could were given the opportunity to attempt the movement to ensure they could safely complete it. Spotters were provided as needed.

### Squatting

Participants were first asked whether they could safely complete the squatting movement. Those who indicated they could proceeded to place each foot on a separate force plate and were instructed to squat as far as they could safely and comfortably go. They were asked to return to a standing position in one fluid motion, keeping their arms extended straight in front of them throughout the movement. Spotters were provided as needed.

### Sit-to-Stand

Participants were first asked whether they could safely stand from a chair without using their arms. Those who indicated they could proceeded with the task. With the standard-height chair (49 cm) positioned just outside the exterior edge of a single force plate, participants were instructed to sit in the chair and place both feet firmly within the force plate’s perimeter, with their arms crossed over their chest. When prompted by the study team, participants rose to a standing position while keeping their arms crossed, paused for one second, and sat back down. When needed, a study team member held the chair steady during each trial to ensure participant safety. Spotters were provided as needed.

### Step-up

Participants were first asked whether they could safely perform the step-up movement. Those who indicated they could proceeded with the task. A standard-height step stool (13 cm) was positioned just outside the edge of a single force plate and placed atop a yoga mat to prevent slipping. For each trial, participants stood directly in front of the step with both feet fully within the force plate perimeter. When prompted by the study team, they stepped up with a single leg, followed by the other, paused at the top, and then stepped down in any manner they chose. Trials were recorded for both left- and right-leg leads, beginning with the left. Spotters were provided as needed.

### Step-down

Participants were first asked whether they could safely perform the step-down movement. Those who indicated they could proceeded with the task. A standard-height step stool (13 cm) was positioned just outside the edge of a single force plate and placed atop a yoga mat to prevent slipping. For each trial, participants stood on the step, facing the force plate. When prompted by the study team, the participant stepped down with a single leg, followed by the other, ensuring both feet landed within the force plate’s perimeter. After each trial, participants stepped back up onto the stool in any manner they chose. Trials were recorded for both left- and right-leg leads, beginning with the left. Spotters were provided as needed.

## Appendix B: Individual ADL timing across all sites

**Table S1.**
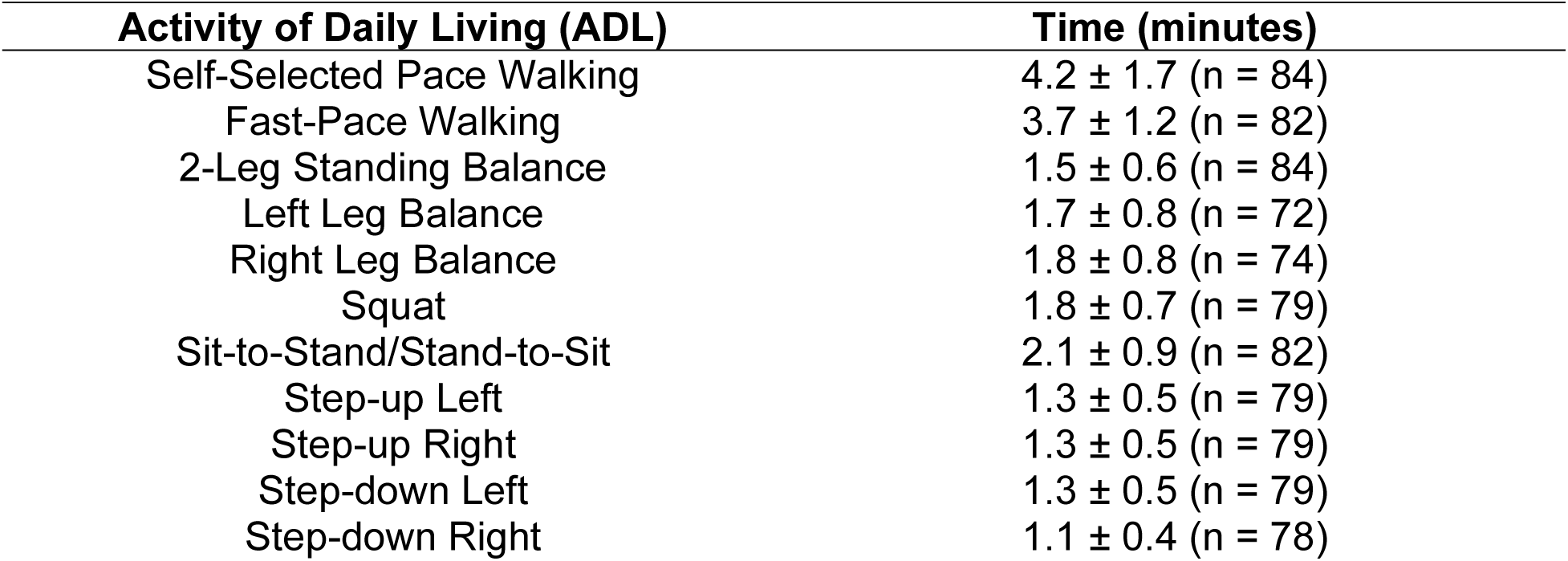
Average time, in minutes, required to complete each ADL across all participants who completed each activity. Missing data reflect non-completion of specific ADL tasks, either by participant choice or for safety reasons.

## Notes

### Competing Interest Statement

The authors have declared no competing interest.

### Author Declarations

The Institutional Review Board of the University of Florida gave ethical approval for this work

